# Clinical Evaluation of a Novel Deep Learning-Based Auto-Segmentation Software: Utility and Potential Pitfalls

**DOI:** 10.64898/2026.01.08.26343652

**Authors:** Ryota Tozuka, Masahide Saito, Masaki Matsuda, Tomoko Akita, Hikaru Nemoto, Takafumi Komiyama, Noriyuki Kadoya, Keiichi Jingu, Hiroshi Onishi

## Abstract

**Background:** Accurate contouring of target volumes and organs at risk is critical for radiotherapy. While deep learning (DL) models offer efficient automation, their generalizability to real-world clinical cases containing anatomical variations and artifacts requires rigorous validation.

**Purpose:** To evaluate the clinical accuracy and robustness of RatoGuide, a novel DL-based auto-segmentation software, using a dataset derived from routine clinical practice including atypical cases.

**Methods:** This single-center retrospective study included 36 patients treated for head and neck, thoracic, abdominal, and pelvic cancers. The cohort was intentionally selected to encompass diverse anatomies and artifacts (e.g., pacemakers, artificial femoral head replacement). Auto-contours generated by RatoGuide were compared with expert-approved manual contours. Performance was evaluated quantitatively using the Dice Similarity Coefficient (DSC) and 95th percentile Hausdorff Distance (HD95), and qualitatively via a 5-point visual assessment scale (higher is better) by four independent reviewers. A score of ≤2 by multiple reviewers was defined as failure.

**Results:** Overall, the mean DSC, HD95, and visual assessment score were 0.79 ± 0.19, 6.35 ± 12.2 mm, and 3.65 ± 0.88, respectively. The mean DSC exceeded 0.8 in 62% (23/37 organ structures) of the evaluated structure types, and a total of 93.5% (315/337) of all contours were considered clinically acceptable based on visual evaluation . However, lower performance was observed in small structures (e.g., optic chiasm) and low-contrast organs (e.g., esophagus).

**Conclusions:** RatoGuide demonstrated favorable performance for major organs across various anatomical regions, consistent with benchmarks reported in the literature. However, performance variability in atypical cases underscores the necessity of rigorous visual verification by experts for clinical implementation.

## Introduction

The accurate contouring of target volumes and organs at risk (OARs) is a critical prerequisite for optimizing treatment plans and evaluating dose distributions in radiation therapy. However, the manual nature of this process, typically conducted by radiation oncologists and medical physicists, is not only time-consuming but also suffers from significant inter-observer variability^1,2^. These limitations can adversely affect tumor control and increase the probability of normal tissue toxicity^3,4^.

Artificial intelligence (AI), particularly deep learning (DL), offers an effective solution to address these challenges and is increasingly used for the auto-segmentation of target volumes and OARs^5–7^. Several studies have demonstrated that AI-assisted segmentation can reduce the clinical workload and decrease both intra- and inter-observer variability^8–11^. In fact, multiple commercial DL-based auto-segmentation software packages are now available and have become integral to modern, high-precision radiotherapy workflows^12–19^.

However, the importance of quality assurance (QA) for DL-based auto-segmentation has been emphasized given that segmentation failures can occur^20,21^. Notably, DL models trained on curated datasets may show reduced generalizability when applied to real-world clinical cases that present with anatomical variations or minor artifacts. Although many reports focus on performance in idealized settings, verifying the applicability to actual clinical workflows is a critical step for software validation. Therefore, evaluating a new DL-based system using clinical datasets that reflect routine practice is essential^22^.

RatoGuide is a novel radiotherapy treatment planning support software that features both auto-segmentation and automated planning capabilities. Although several reports have evaluated its automated planning functions, its auto-segmentation performance has not yet been comprehensively assessed^23–25^. To address this, we investigated the clinical applicability of RatoGuide by comparing its auto-contours with manual contouring using clinical cases derived from routine practice.

## Materials and Methods

### Patient Dataset and Image Acquisition

This study was approved by the institutional review board of the University of Yamanashi. The receipt number was CS0010. This retrospective study included 36 patients treated at our institution between November 2018 and March 2025. To evaluate the software’s generalizability to routine clinical practice, cases involving the head and neck, thorax, abdomen, and male pelvis regions were selected from our database. The cohort was curated to encompass diverse tumor types and anatomical variations, including atypical cases, to assess the software’s robustness in real-world scenarios. Patient characteristics are summarized in Table 1. Planning CT scans were acquired using an Aquilion LB (Canon Medical Systems, Otawara, Japan). All patients underwent non-contrast planning CT scanning with the following parameters: 120 kV, 250 mA, and 2-mm slice thickness. Metal artifact reduction (MAR) was applied in selected cases. Necessary pre-treatment procedures, such as bladder filling for prostate cancer irradiation, were performed as clinically indicated.

**Table 1.** Clinical characteristics of the cases included in this study.

|  |  |  |
| --- | --- | --- |
| Head and Neck |  |  |
|  | Sinus Ca. | 2 |
|  | Nasopharyngeal Ca. | 2 |
|  | Oropharyngeal Ca. | 2 |
|  | Hypopharyngeal Ca. | 2 |
|  | Oral Ca. | 2 |
| Thorax |  |  |
|  | Lung Ca. | 8 |
|  | Early-stage | ✓ (2) |
|  | Advanced-stage | ✓ (2) |
|  | with a pacemaker | ✓ (2) |
|  | Unilateral atelectasis | ✓ (2) |
| Abdomen |  |  |
|  | Liver Ca. | 2 |
|  | Adrenal Ca. | 2 |
|  | Pancreatic Ca. | 2 |
|  | Gastric MALT lymphoma | 2 |
| Male Pelvis |  |  |
|  | Prostate Ca. | 10 |
|  | without SpaceOAR | ✓ (2) |
|  | with SpaceOAR | ✓ (2) |
|  | poor bladder filling (<100cc) | ✓ (2) |
|  | Artificial femoral head replacement | ✓ (2) |
|  | Excessive rectal gas | ✓ (2) |

The clinical contours used for treatment served as the ground truth (GT) for this study. Delineation was performed on either the Pinnacle (Philips Radiation Oncology Systems, Fitchburg, WI, USA) or RayStation (RaySearch Laboratories, Stockholm, Sweden) treatment planning systems, based on standard delineation guidelines^26–30^. Where appropriate, fused MRI or contrast-enhanced CT (CECT) images were used to assist in accurate delineation. Initially drawn by a radiation oncologist or a medical physicist, all contours were subsequently reviewed, revised if necessary, and formally approved by a senior radiation oncologist with more than 7 years of experience.

To ensure fair evaluation for longitudinally extensive organs (e.g., the esophagus and spinal cord), the craniocaudal extent of the AI contours was adjusted. Since manual contours are often limited to the clinically relevant region, AI contours extending beyond the manual ground truth (GT) were cropped to match the GT limits in the superior-inferior direction. Conversely, if the AI contour was shorter than the GT, no adjustment was made to account for under-segmentation^31^.

### Generation of AI-based Contours

RatoGuide (Version 1.6.1.7, research version) is a deep learning-based automatic segmentation software utilizing the UNesT model architecture^32^. Specific details regarding the training dataset and pre-processing algorithms are proprietary to the manufacturer and were not disclosed.

In the experimental workflow, planning CT datasets in DICOM format were imported into RatoGuide using external storage devices, as the system operated in a standalone environment isolated from the institutional network. Upon selecting the target structures, the auto-segmentation process was executed. The generated contours were subsequently exported as RT-Structure sets to RayStation for evaluation.

### Quantitative and Qualitative Evaluations

To evaluate the auto-contours generated by RatoGuide, we employed two types of assessments: quantitative and qualitative evaluations.

For the quantitative evaluation, the Dice Similarity Coefficient (DSC) and the 95th percentile Hausdorff Distance (HD95) were employed. The DSC is a metric that indicates the degree of overlap between two segmented volumes, A and B, and is defined by the following equation:

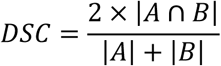

where |A| and |B| represent the volumes of the two segments. Generally, a DSC value of 0.8 or higher is considered to indicate good agreement^33^.

Furthermore, the HD95 was utilized to evaluate the boundary accuracy. The Hausdorff Distance measures the maximum distance from a point in one set to the nearest point in the other. To eliminate the impact of outliers, the 95th percentile of these distances is used instead of the maximum. It is defined based on the directed Hausdorff distance ℎ(*A*, *B*):

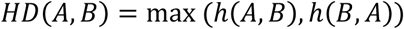

where ℎ(*A*, *B*) = *max*_*a*∈*A*_*min*_*b*∈*B*_‖*a* − *b*‖. For HD95, the 95th percentile is calculated in place of the maximum operation.

While DSC is a widely used metric, it has known limitations: it can be influenced by volume size and correlates poorly with clinical acceptability^34,35^. Moreover, considering that the manual contours used as the ground truth inherently contain inter-observer variability, relying solely on quantitative metrics may not yield a comprehensive assessment. Therefore, a qualitative evaluation was performed by four independent reviewers: two experienced radiation oncologists and two medical physicists. The AI contours were visually reviewed and scored on a 5-point scale based on the criteria outlined in Table 2. A score of 3 was defined as the minimum threshold for clinical acceptability. An organ was classified as a "failure" if two or more evaluators assigned a score of 2 or lower (i.e., judged the AI segmentation to be clinically unacceptable).

**Table 2.** Qualitative Evaluation Criteria.

| Score | Criteria |
| --- | --- |
| 5 | **Acceptable**; clinically usable without modification |
| 4 | **Acceptable**; clinically usable despite minor deviations |
| 3 | **Acceptable**; requires revisions for clinical use |
| 2 | **Unacceptable**; requires complete redrawing. |
| 1 | **Unacceptable**; organ not recognized |

## Results

Table 3 summarizes the mean (± standard deviation) number of CT slices and inference time for each anatomical region. The overall mean inference time was 91.4 ± 13.4 s. The maximum inference time observed in this dataset was 139 s, corresponding to a head and neck case comprising 263 CT slices.

**Table 3.** Mean number of CT slices and inference time by anatomical region.

|  | Head and Neck | Thorax | Abdomen | Male pelvis |
| --- | --- | --- | --- | --- |
| CT images | 207.3 ± 27.0 | 218.3 ± 26.8 | 173.6 ± 17.9 | 173.7 ± 22.5 |
| Auto-segmentation time [sec] | 93.7 ± 20.4 | 92.8 ± 13.3 | 87.9 ± 9.19 | 90.8 ± 16.9 |
| (Ave ± SD) |  |  |  |  |

Figures 1 and 2 present the DSC and HD95 results for all organs across each region. The mean DSC values were 0.73 ± 0.19, 0.79 ± 0.22, 0.90 ± 0.05, and 0.84 ± 0.16 for the head and neck, thorax, abdomen, and pelvis, respectively. For the same regions, the mean HD95 values were 4.40 ± 3.71 mm, 13.60 ± 25.40 mm, 4.57 ± 3.29 mm, and 5.27 ± 5.70 mm, respectively. Furthermore, the mean DSC exceeded 0.8 in 62% (23/37 structure types) of the evaluated contours.

**Figure 1.**
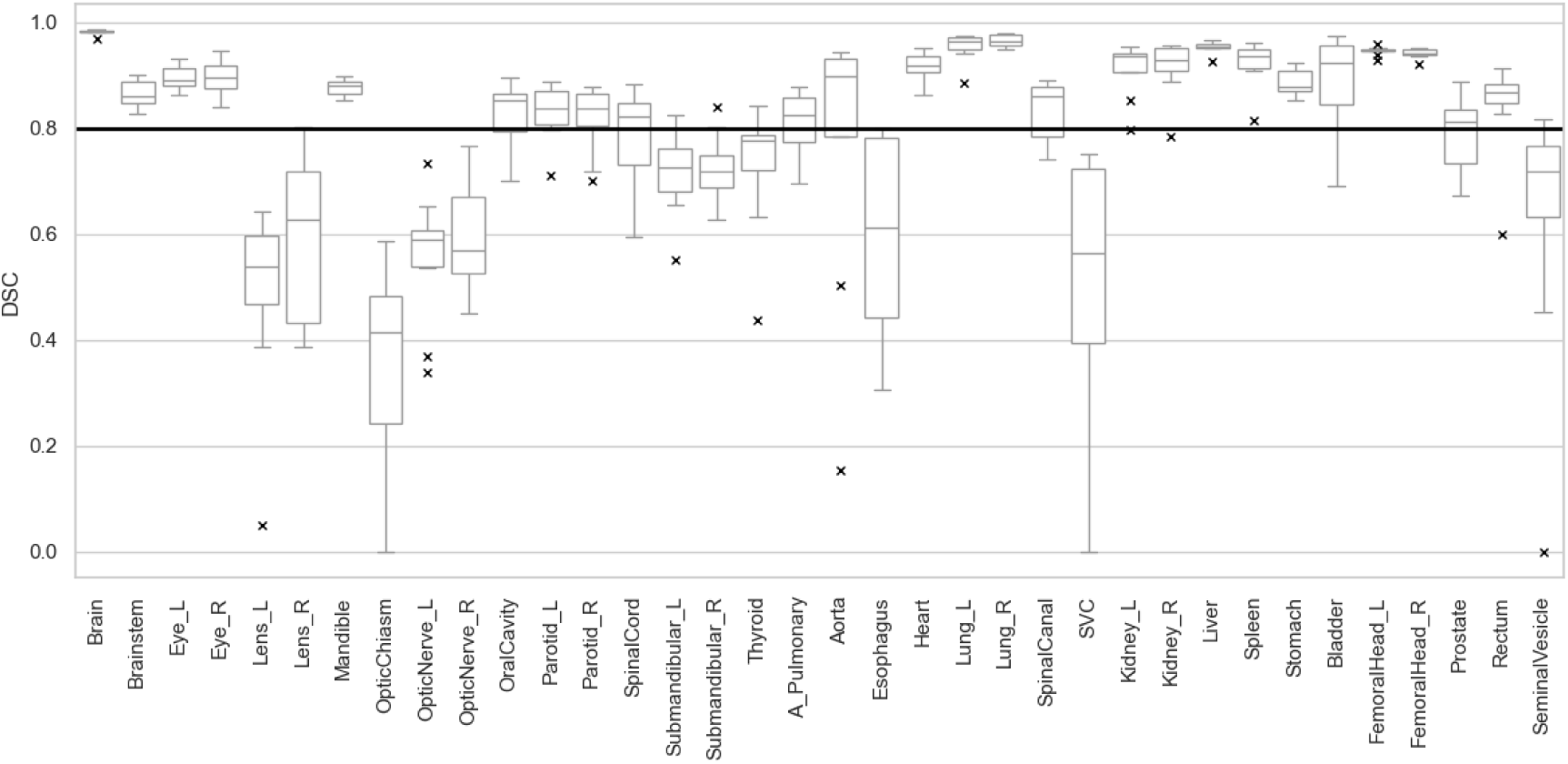
Box plots of Dice Similarity Coefficients (DSC) for all evaluated structures.

**Figure 2.**
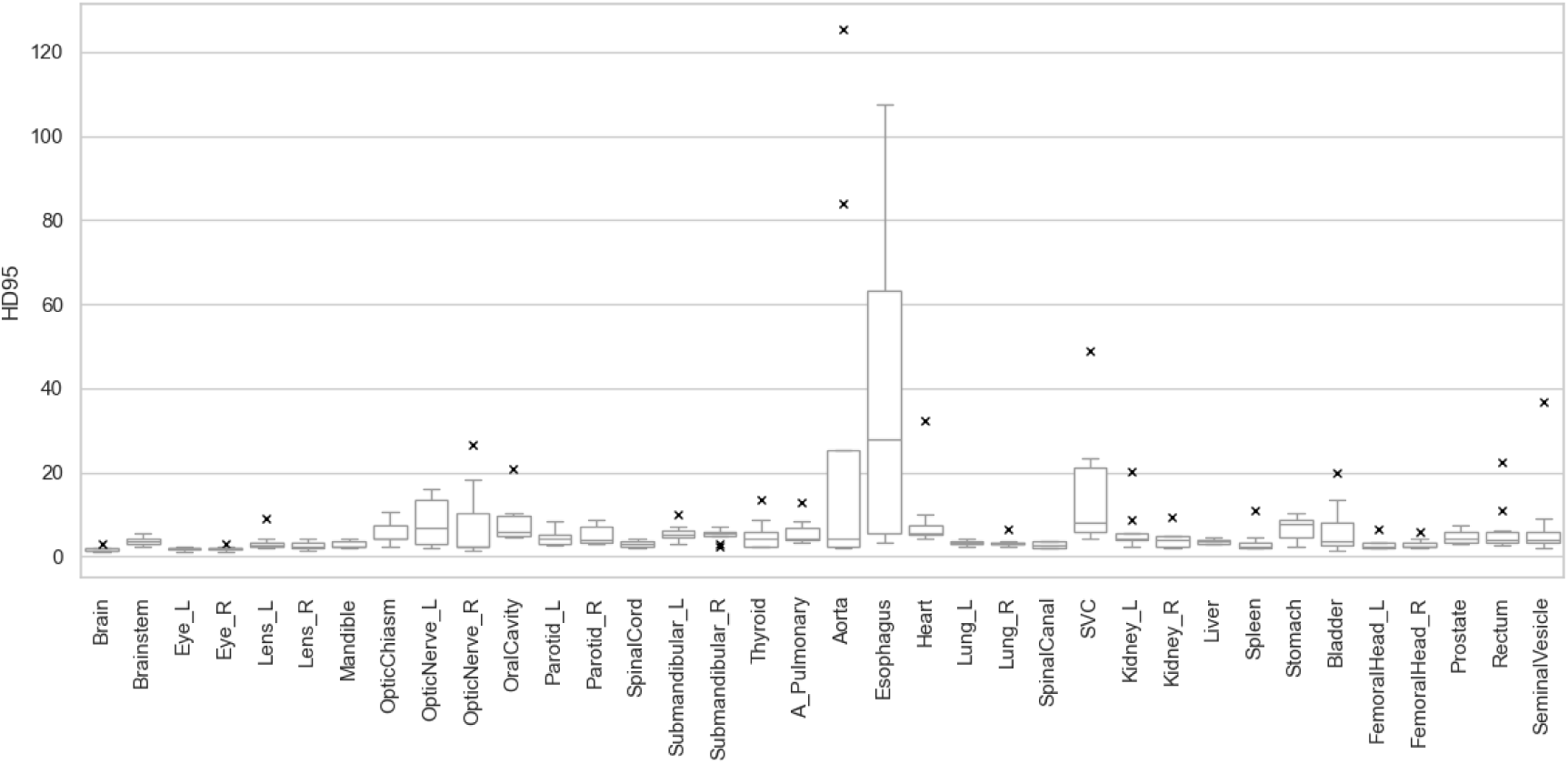
Box plots of the 95th percentile Hausdorff Distance (HD95) for all evaluated structures..

Table 4 presents the results of the visual assessment performed by four independent reviewers. The mean visual scores were 3.76 ± 0.83, 3.47 ± 1.03, 3.50 ± 0.77, and 3.65 ± 0.85 for the head and neck, thorax, abdomen, and pelvis, respectively. The overall failure rate, defined as contours receiving a score of 2 or lower by at least two reviewers, was 6.5%.

**Table 4.**
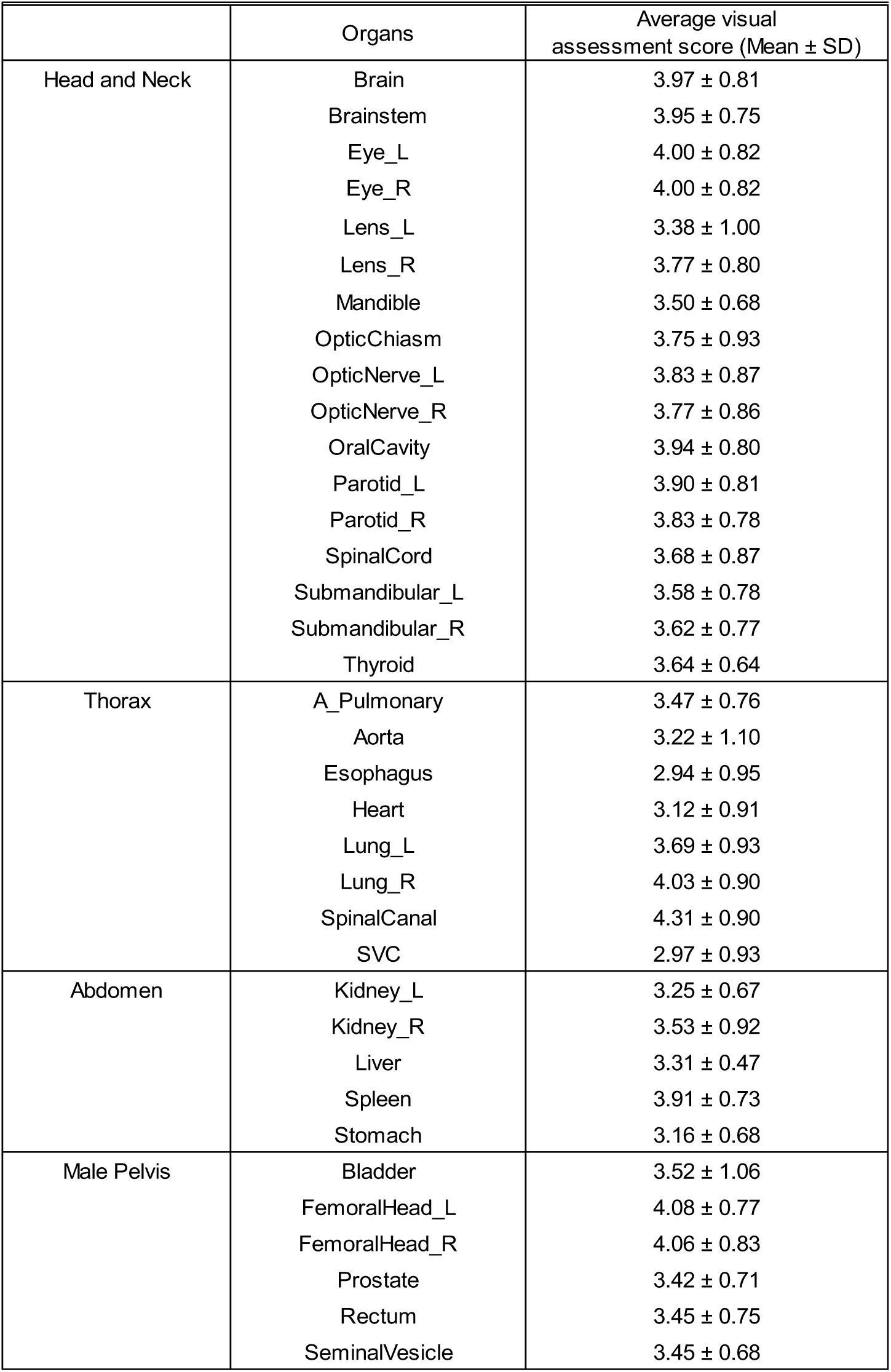
Results of visual assessment by four independent reviewers.

| | Organs | Average visual assessment score (Mean ± SD) |
| --- | --- | --- |
| Head and Neck | Brain | 3.97 ± 0.81 |
| | Brainstem | 3.95 ± 0.75 |
| | Eye_L | 4.00 ± 0.82 |
| | Eye_R | 4.00 ± 0.82 |
| | Lens_L | 3.38 ± 1.00 |
| | Lens_R | 3.77 ± 0.80 |
| | Mandible | 3.50 ± 0.68 |
| | OpticChiasm | 3.75 ± 0.93 |
| | OpticNerve_L | 3.83 ± 0.87 |
| | OpticNerve_R | 3.77 ± 0.86 |
| | OralCavity | 3.94 ± 0.80 |
| | Parotid_L | 3.90 ± 0.81 |
| | Parotid_R | 3.83 ± 0.78 |
| | SpinalCord | 3.68 ± 0.87 |
| | Submandibular_L | 3.58 ± 0.78 |
| | Submandibular_R | 3.62 ± 0.77 |
| | Thyroid | 3.64 ± 0.64 |
| Thorax | A_Pulmonary | 3.47 ± 0.76 |
| | Aorta | 3.22 ± 1.10 |
| | Esophagus | 2.94 ± 0.95 |
| | Heart | 3.12 ± 0.91 |
| | Lung_L | 3.69 ± 0.93 |
| | Lung_R | 4.03 ± 0.90 |
| | SpinalCanal | 4.31 ± 0.90 |
| | SVC | 2.97 ± 0.93 |
| Abdomen | Kidney_L | 3.25 ± 0.67 |
| | Kidney_R | 3.53 ± 0.92 |
| | Liver | 3.31 ± 0.47 |
| | Spleen | 3.91 ± 0.73 |
| | Stomach | 3.16 ± 0.68 |
| Male Pelvis | Bladder | 3.52 ± 1.06 |
| | FemoralHead_L | 4.08 ± 0.77 |
| | FemoralHead_R | 4.06 ± 0.83 |
| | Prostate | 3.42 ± 0.71 |
| | Rectum | 3.45 ± 0.75 |
| | SeminalVesicle | 3.45 ± 0.68 |

## Discussion

AI-based automated segmentation is increasingly integrated into clinical practice to enhance workflow efficiency and standardize delineation. In this study, we evaluated the accuracy of RatoGuide, a newly released deep learning-based auto-segmentation software, using a dataset encompassing clinical cases derived from routine practice. Overall, the mean DSC, HD95 and visual assessment score were 0.79 ± 0.19, 6.35 ± 12.2 mm and 3.65 ± 0.88, respectively, with 6.5% of cases classified as failures. These DSC and HD95 values align with the “moderate agreement” benchmarks reported in the literature^14,36^. Crucially, this study evaluated not only typical cases but also challenging ones, such as patients with artificial femoral head replacements. Evaluating performance in such a heterogeneous population is clinically vital. It is noteworthy that the software achieved these accuracy levels despite these real-world complexities^37^. The auto-segmentation accuracy of RatoGuide is broadly consistent with previously published data. Doolan et al. evaluated five commercial auto-segmentation software packages and reported median volumetric DSCs ranging from 0.82 to 0.88^14^. For comparison, the median DSC for RatoGuide in this study was 0.85. Although a direct comparison is challenging due to differences in patient cohorts and target organs, this result falls within the range of previously reported values.

### Head and neck cases

In the head and neck region, moderate to good DSCs were observed for most structures. Specifically, structures other than the lens, optic nerves, and optic chiasm achieved visual assessment scores of > 3.5 with no failures. In contrast, the lens, optic nerves, and optic chiasm showed lower performance in terms of both DSC and HD95. Notably, the left lens was rated as a failure in only one case (Figure 3). The reduced accuracy observed in these structures can be attributed to their small volumes, as the DSC is inherently sensitive to size and tends to penalize smaller structures more heavily^13,14,38^. Furthermore, given that the optic nerves and chiasm are generally exhibit low soft-tissue contrast on CT images and are known challenges for CT-based auto-segmentation, these results align with well-known limitations in the field^39,40^.

**Figure 3.**
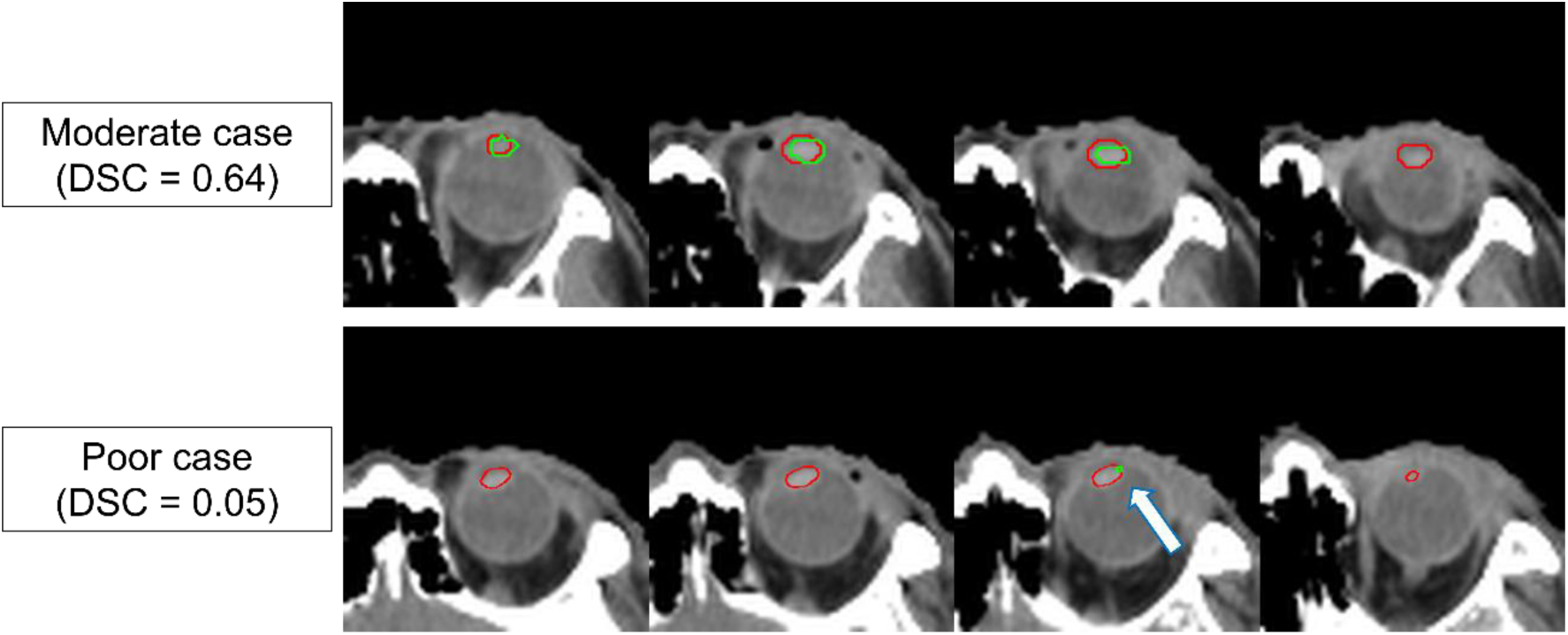
Representative examples of lens segmentation. Green lines indicate auto-segmentation, and red lines indicate ground truth. (a) A moderate case. (b) A poor case. As indicated by the arrow, the AI significantly under-segmented the lens.

### Thorax cases

In the thoracic region, moderate to good DSCs were achieved for most structures, with the exception of the esophagus and superior vena cava (SVC). These organs present small cross-sectional areas in the axial plane compared to larger organs such as the lungs and heart. The esophagus, in particular, is widely regarded as one of the most challenging organs for auto-segmentation due to its low contrast against surrounding soft tissues, tortuous path, and tendency to be compressed^13,19^. In our study, 4 out of 8 cases were rated as failures; a result consistent with the inherent difficulty of delineating this organ on non-contrast planning CTs. Failures in other organs occurred in cases with significant anatomical distortion (e.g., atelectasis). Generally, pathological conditions such as atelectasis are prone to auto-segmentation failure because such cases are insufficiently represented in training datasets to allow for model generalization; and RatoGuide was no exception to this trend. Conversely, high accuracy was unexpectedly maintained in cases with implanted pacemakers, even though training data for such artifacts was also expected to be limited (Figure 4). This indicates that the impact of artifacts on auto-segmentation quality can be variable. While the system shows potential in specific scenarios, this unpredictability highlights that rigorous visual verification is essential when applying auto-segmentation to atypical cases.

**Figure 4.**
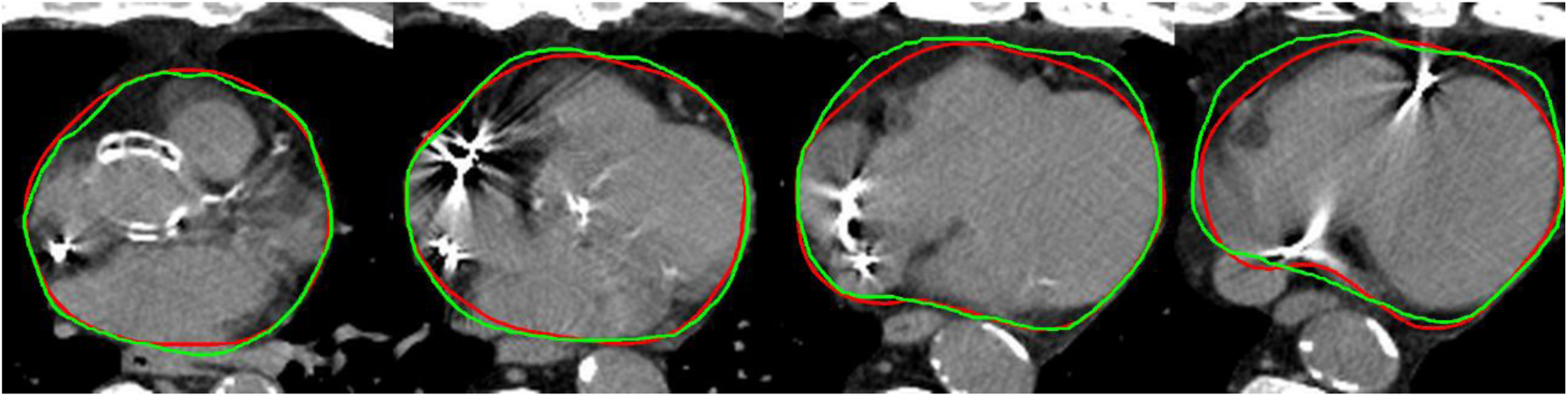
Segmentation result in a patient with an implanted pacemaker. Despite metal artifacts caused by the pacemaker leads, the target structure was successfully delineated (green) relative to the ground truth (red).

### Abdomen cases

In the abdominal region, good accuracy was achieved with DSC exceeding 0.8 for all structures. However, these quantitative metrics did not consistently align with the visual assessments; failures were observed in the kidneys and stomach in some cases. Of particular concern was a failure in kidney segmentation in a case that appeared anatomically normal to the naked eye (Figure 5). Typically, auto-segmentation failures are attributable to anomalous features that are underrepresented in the training dataset, making them somewhat predictable. In contrast, the error observed here was "unanticipated," occurring in the absence of obvious anatomical triggers. Such unpredictable failures in seemingly normal cases are a known risk in AI-based segmentation, extending beyond specific anatomical regions. For instance, Temple et al. similarly reported two cases of such unanticipated errors in the parotid glands, highlighting that this phenomenon is a general challenge for deep learning models rather than an organ-specific issue^22^. These errors pose a high risk of automation bias, as users do not expect the system to fail in routine cases. These findings underscore the critical need for radiation oncologists and medical physicists to rigorously evaluate AI-generated contours qualitatively, even for cases that appear straightforward, rather than relying solely on quantitative metrics.

**Figure 5.**
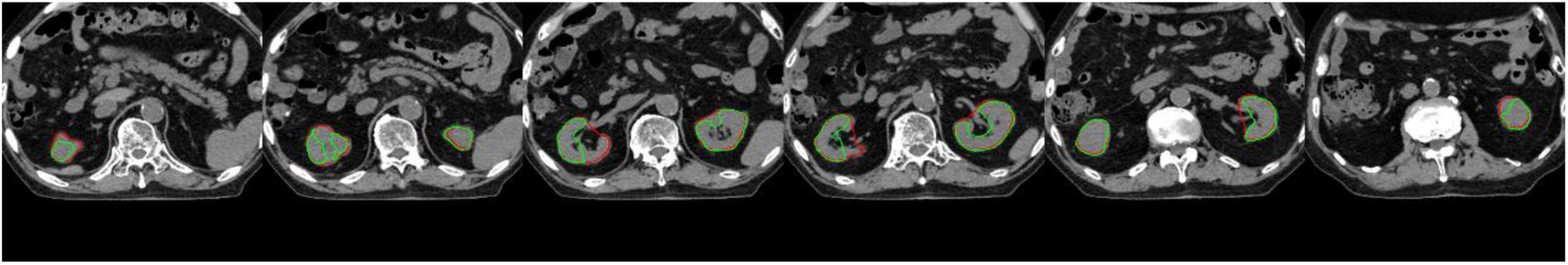
Unexpected segmentation failure in a visually normal case. Example of failure in the kidney region, where green lines indicate auto-segmentation and red lines indicate ground truth. Unlike complex cases where errors are anticipated, failures in such typical cases pose a risk of automation bias.

### Male pelvis cases

In the male pelvic region, good accuracy was observed for all structures with the exception of the seminal vesicles. The lower DSCs for the seminal vesicles are consistent with previous reports and are primarily attributed to the low soft-tissue contrast between the organ and surrounding tissues, which results in indistinct boundaries on CT images^41^. While incorporating MRI as an input (multi-modal approach) could potentially resolve this issue, the current version of RatoGuide does not support such functionality^42^. Regarding atypical anatomy, results stood in contrast to the pacemaker cases observed in the thorax. In cases involving artificial femoral head replacement, severe metal artifacts degraded image quality across the pelvis (Figure 6). This led to gross segmentation failures in multiple organs. Notably, failures persisted even in a case where a MAR was applied. This suggests that while the model may tolerate localized artifacts (like pacemakers), wide-ranging image degradation remains a critical limitation that cannot be easily mitigated by standard reconstruction techniques.

**Figure 6.**
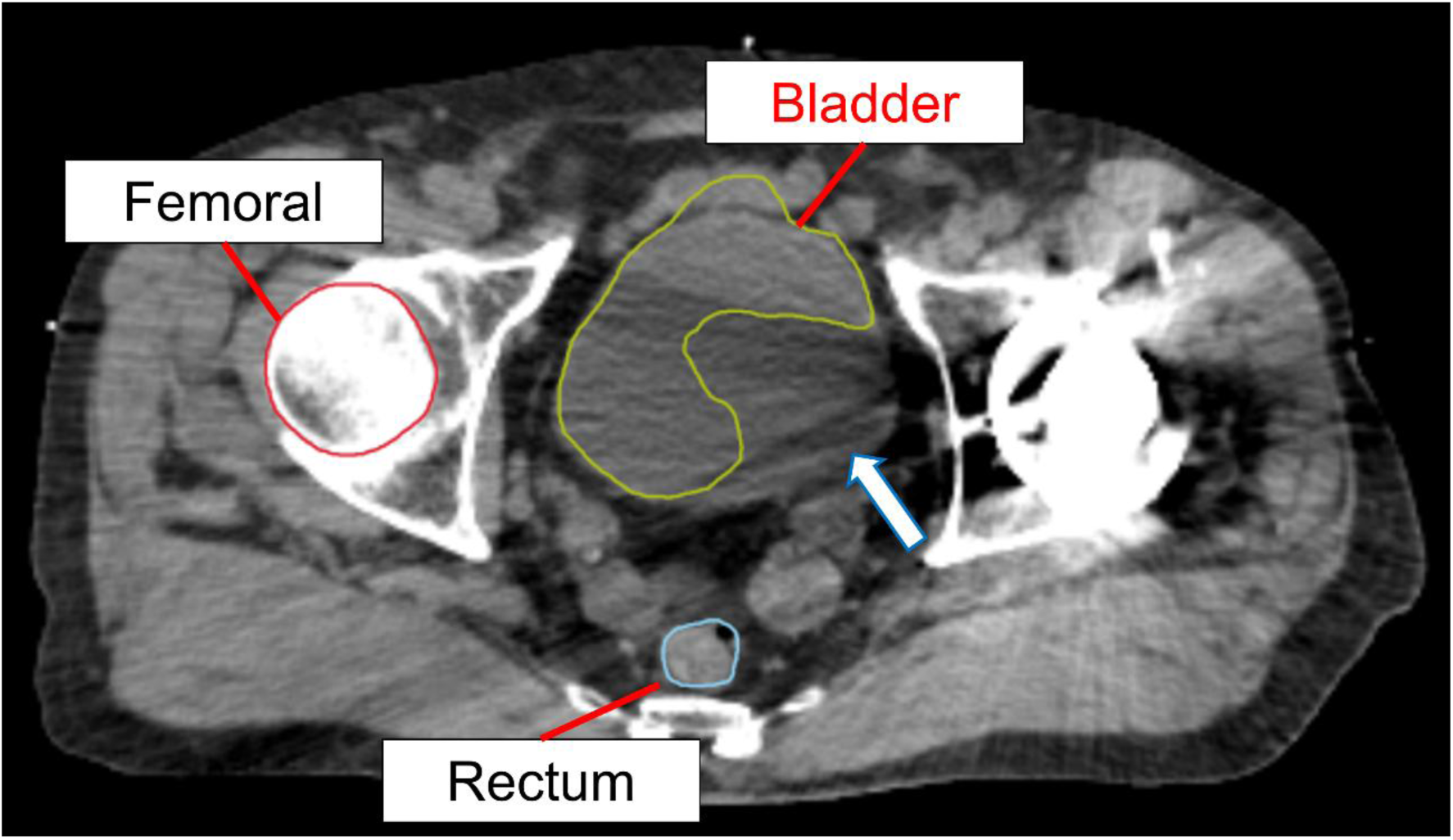
Failure in bladder segmentation due to metal artifacts. Case following artificial femoral head replacement. Artifacts from the metal degraded the contrast, leading to incomplete delineation of the bladder as indicated by the arrow.

### Limitation

This study has several limitations. First, our evaluation focused exclusively on RatoGuide without direct comparison to other commercial solutions. Future research should aim to perform a comparative analysis using multiple software packages to ensure a more robust evaluation. Second, the patient cohort was small and limited to a single institution. Since institution-specific parameters, such as CT acquisition protocols, can influence auto-segmentation accuracy, larger-scale, multi-institutional studies are necessary to validate these findings. Third, the ground truth contours used in this study were derived from clinical practice and thus contain inherent inter-observer variability. While reviewed by experts, it should be acknowledged that the GT itself may not be the absolute gold standard.

## Conclusion

In conclusion, RatoGuide, a novel deep learning-based auto-segmentation software, demonstrated favorable performance for major organs across various anatomical regions. Nevertheless, visual verification by radiation oncologists and medical physicists remains essential for clinical implementation, particularly in atypical cases.

## Author Contributions

RT, NK contributed to the conception and design of this study. MS, MM, TA and HN conducted the primary analysis. RT mainly drafted the manuscript. NK, TK, KJ, and HO reviewed the manuscript. All authors read and approved the final manuscript.

## Data Availability

All data produced in the present study are available upon reasonable request to the authors.

## Acknowledgements

We partially used Gemini for English language editing, but the final text was fully reviewed and corrected by the authors.

This study was approved by the institutional review board of the University of Yamanashi. The receipt number was CS0010.

## References

1. Li XA, Tai A, Arthur DW, et al. Variability of target and normal structure delineation for breast cancer radiotherapy: an RTOG Multi-Institutional and Multiobserver Study. Int J Radiat Oncol Biol Phys. Mar 01 2009;73(3):944–51. doi:10.1016/j.ijrobp.2008.10.034

2. Vinod SK, Jameson MG, Min M, Holloway LC. Uncertainties in volume delineation in radiation oncology: A systematic review and recommendations for future studies. Radiother Oncol. Nov 2016;121(2):169–179. doi:10.1016/j.radonc.2016.09.009

3. Nelms BE, Tomé WA, Robinson G, Wheeler J. Variations in the contouring of organs at risk: test case from a patient with oropharyngeal cancer. Int J Radiat Oncol Biol Phys. Jan 01 2012;82(1):368–78. doi:10.1016/j.ijrobp.2010.10.019

4. Jaikuna T, Osorio EV, Azria D, et al. Contouring variation affects estimates of normal tissue complication probability for breast fibrosis after radiotherapy. Breast. Dec 2023;72:103578. doi:10.1016/j.breast.2023.103578

5. Erdur AC, Rusche D, Scholz D, et al. Deep learning for autosegmentation for radiotherapy treatment planning: State-of-the-art and novel perspectives. Strahlenther Onkol. Mar 2025;201(3):236–254. doi:10.1007/s00066-024-02262-2

6. Tozuka R, Kadoya N, Yasunaga A, et al. Fractal-driven self-supervised learning enhances early-stage lung cancer GTV segmentation: a novel transfer learning framework. Jpn J Radiol. Sep 15 2025; doi:10.1007/s11604-025-01865-8

7. Marin T, Zhuo Y, Lahoud RM, et al. Deep learning-based GTV contouring modeling inter- and intra- observer variability in sarcomas. Radiother Oncol. Feb 2022;167:269–276. doi:10.1016/j.radonc.2021.09.034

8. Han Z, Wang Y, Wang W, et al. Artificial intelligence-assisted delineation for postoperative radiotherapy in patients with lung cancer: a prospective, multi-center, cohort study. Front Oncol. 2024;14:1388297. doi:10.3389/fonc.2024.1388297

9. Choi MS, Chang JS, Kim K, et al. Assessment of deep learning-based auto-contouring on interobserver consistency in target volume and organs-at-risk delineation for breast cancer: Implications for RTQA program in a multi-institutional study. Breast. Feb 2024;73:103599. doi:10.1016/j.breast.2023.103599

10. Pang EPP, Tan HQ, Wang F, et al. Multicentre evaluation of deep learning CT autosegmentation of the head and neck region for radiotherapy. NPJ Digit Med. May 27 2025;8(1):312. doi:10.1038/s41746-025-01624-z

11. Ginn JS, Gay HA, Hilliard J, et al. A clinical and time savings evaluation of a deep learning automatic contouring algorithm. Med Dosim. 2023 Spring 2023;48(1):55–60. doi:10.1016/j.meddos.2022.11.001

12. Rusanov B, Ebert MA, Sabet M, et al. Guidance on selecting and evaluating AI auto-segmentation systems in clinical radiotherapy: insights from a six-vendor analysis. Phys Eng Sci Med. Mar 2025;48(1):301–316. doi:10.1007/s13246-024-01513-x

13. Kim YW, Biggs S, Claridge Mackonis E. Investigation on performance of multiple AI-based auto-contouring systems in organs at risks (OARs) delineation. Phys Eng Sci Med. Sep 2024;47(3):1123–1140. doi:10.1007/s13246-024-01434-9

14. Doolan PJ, Charalambous S, Roussakis Y, et al. A clinical evaluation of the performance of five commercial artificial intelligence contouring systems for radiotherapy. Front Oncol. 2023;13:1213068. doi:10.3389/fonc.2023.1213068

15. Fan M, Wang T, Lei Y, et al. Evaluation and failure analysis of four commercial deep learning-based autosegmentation software for abdominal organs at risk. J Appl Clin Med Phys. Apr 2025;26(4):e70010. doi:10.1002/acm2.70010

16. Meyer C, Huger S, Bruand M, et al. Artificial intelligence contouring in radiotherapy for organs-at-risk and lymph node areas. Radiat Oncol. Nov 21 2024;19(1):168. doi:10.1186/s13014-024-02554-y

17. Choi SH, Park JW, Cho Y, Yang G, Yoon HI. Automated Organ Segmentation for Radiation Therapy: A Comparative Analysis of AI-Based Tools Versus Manual Contouring in Korean Cancer Patients. Cancers (Basel). Oct 30 2024;16(21)doi:10.3390/cancers16213670

18. Rong Y, Chen Q, Fu Y, et al. NRG Oncology Assessment of Artificial Intelligence Deep Learning-Based Auto-segmentation for Radiation Therapy: Current Developments, Clinical Considerations, and Future Directions. Int J Radiat Oncol Biol Phys. May 01 2024;119(1):261–280. doi:10.1016/j.ijrobp.2023.10.033

19. Yuan L, Chen Q, Al-Hallaq H, et al. Quantitative Evaluation of Artificial Intelligence-Based Organ Segmentation Across Multiple Anatomic Sites Using 8 Commercial Software Platforms. Pract Radiat Oncol. Aug 23 2025;doi:10.1016/j.prro.2025.06.012

20. Cardenas CE, Yang J, Anderson BM, Court LE, Brock KB. Advances in Auto-Segmentation. Semin Radiat Oncol. Jul 2019;29(3):185–197. doi:10.1016/j.semradonc.2019.02.001

21. Vandewinckele L, Claessens M, Dinkla A, et al. Overview of artificial intelligence-based applications in radiotherapy: Recommendations for implementation and quality assurance. Radiother Oncol. Dec 2020;153:55–66. doi:10.1016/j.radonc.2020.09.008

22. Temple SWP, Rowbottom CG. Gross failure rates and failure modes for a commercial AI-based auto-segmentation algorithm in head and neck cancer patients. J Appl Clin Med Phys. Jun 2024;25(6):e14273. doi:10.1002/acm2.14273

23. Kadoya N, Kimura Y, Tozuka R, et al. Evaluation of deep learning-based deliverable VMAT plan generated by prototype software for automated planning for prostate cancer patients. J Radiat Res. Sep 22 2023;64(5):842–849. doi:10.1093/jrr/rrad058

24. Nemoto H, Saito M, Kadoya N, et al. Evaluation of deep learning-based automated radiotherapy planning for early-stage lung cancer using SBRT-VMAT: A comparison with manual planning. J Appl Clin Med Phys. Oct 2025;26(10):e70291. doi:10.1002/acm2.70291

25. Xiao Y, Tanaka S, Kadoya N, et al. Evaluation of deliverable artificial intelligence-based automated volumetric arc radiation therapy planning for whole pelvic radiation in gynecologic cancer. Sci Rep. Apr 30 2025;15(1):15219. doi:10.1038/s41598-025-99717-y

26. Brouwer CL, Steenbakkers RJ, Bourhis J, et al. CT-based delineation of organs at risk in the head and neck region: DAHANCA, EORTC, GORTEC, HKNPCSG, NCIC CTG, NCRI, NRG Oncology and TROG consensus guidelines. Radiother Oncol. Oct 2015;117(1):83–90. doi:10.1016/j.radonc.2015.07.041

27. Kong FM, Ritter T, Quint DJ, et al. Consideration of dose limits for organs at risk of thoracic radiotherapy: atlas for lung, proximal bronchial tree, esophagus, spinal cord, ribs, and brachial plexus. Int J Radiat Oncol Biol Phys. Dec 01 2011;81(5):1442–57. doi:10.1016/j.ijrobp.2010.07.1977

28. Feng M, Moran JM, Koelling T, et al. Development and validation of a heart atlas to study cardiac exposure to radiation following treatment for breast cancer. Int J Radiat Oncol Biol Phys. Jan 01 2011;79(1):10–8. doi:10.1016/j.ijrobp.2009.10.058

29. Jabbour SK, Hashem SA, Bosch W, et al. Upper abdominal normal organ contouring guidelines and atlas: a Radiation Therapy Oncology Group consensus. Pract Radiat Oncol. 2014;4(2):82–89. doi:10.1016/j.prro.2013.06.004

30. Gay HA, Barthold HJ, O’Meara E, et al. Pelvic normal tissue contouring guidelines for radiation therapy: a Radiation Therapy Oncology Group consensus panel atlas. Int J Radiat Oncol Biol Phys. Jul 01 2012;83(3):e353–62. doi:10.1016/j.ijrobp.2012.01.023

31. Finnegan RN, Quinn A, Horsley P, et al. Geometric and dosimetric evaluation of a commercial AI auto-contouring tool on multiple anatomical sites in CT scans. J Appl Clin Med Phys. Jun 2025;26(6):e70067. doi:10.1002/acm2.70067

32. Yu X, Yang Q, Zhou Y, et al. Unest: local spatial representation learning with hierarchical transformer for efficient medical segmentation. Medical Image Analysis. 2023;90:102939.

33. Brock KK, Mutic S, McNutt TR, Li H, Kessler ML. Use of image registration and fusion algorithms and techniques in radiotherapy: Report of the AAPM Radiation Therapy Committee Task Group No. 132. Med Phys. Jul 2017;44(7):e43–e76. doi:10.1002/mp.12256

34. Sherer MV, Lin D, Elguindi S, et al. Metrics to evaluate the performance of auto-segmentation for radiation treatment planning: A critical review. Radiother Oncol. Jul 2021;160:185–191. doi:10.1016/j.radonc.2021.05.003

35. Mackay K, Bernstein D, Glocker B, Kamnitsas K, Taylor A. A Review of the Metrics Used to Assess Auto-Contouring Systems in Radiotherapy. Clin Oncol (R Coll Radiol). Jun 2023;35(6):354–369. doi:10.1016/j.clon.2023.01.016

36. Brouwer CL, Steenbakkers RJ, van den Heuvel E, et al. 3D Variation in delineation of head and neck organs at risk. Radiat Oncol. Mar 13 2012;7:32. doi:10.1186/1748-717X-7-32

37. Kanwar A, Merz B, Claunch C, Rana S, Hung A, Thompson RF. Stress-testing pelvic autosegmentation algorithms using anatomical edge cases. Phys Imaging Radiat Oncol. Jan 2023;25:100413. doi:10.1016/j.phro.2023.100413

38. Ibragimov B, Xing L. Segmentation of organs-at-risks in head and neck CT images using convolutional neural networks. Med Phys. Feb 2017;44(2):547–557. doi:10.1002/mp.12045

39. Liu Y, Lei Y, Fu Y, et al. Head and neck multi-organ auto-segmentation on CT images aided by synthetic MRI. Med Phys. Sep 2020;47(9):4294–4302. doi:10.1002/mp.14378

40. Mlynarski P, Delingette H, Alghamdi H, Bondiau PY, Ayache N. Anatomically consistent CNN-based segmentation of organs-at-risk in cranial radiotherapy. J Med Imaging (Bellingham). Jan 2020;7(1):014502. doi:10.1117/1.JMI.7.1.014502

41. Miura H, Ishihara S, Kenjo M, Nakao M, Ozawa S, Kagemoto M. Evaluation of the accuracy of automated segmentation based on deep learning for prostate cancer patients. Med Dosim. 2025 Spring 2025;50(1):91–95. doi:10.1016/j.meddos.2024.09.002

42. Elguindi S, Zelefsky MJ, Jiang J, et al. Deep learning-based auto-segmentation of targets and organs-at-risk for magnetic resonance imaging only planning of prostate radiotherapy. Phys Imaging Radiat Oncol. Oct 2019;12:80–86. doi:10.1016/j.phro.2019.11.006

